# Head-to-head comparison of [^18^F]florbetapir and [^18^F]FDG PET for the early detection of amyloidosis in systemic amyloidosis and plasma cell dyscrasias

**DOI:** 10.1101/2024.08.19.24312210

**Authors:** Yanyan Kong, Lei Cao, Boyan He, Zhongwen Zhou, Minmin Zhang, Qian Zhang, Qian Wang, Wei Wang, Haoxiang Zhu, Jianfei Xiao, Axel Rominger, Yihui Guan, Haibo Tan, Ruiqing Ni

## Abstract

**Purpose:** Amyloidosis is underdiagnosed in light-chain amyloidosis (AL) and hereditary transthyretin amyloidosis (hATTR), as well as plasma cell dyscrasias (PCD). We aimed to investigate the utility of [^18^F]florbetapir (FBP) and [^18^F]fluorodeoxyglucose (FDG) positron emission tomography (PET) for the early detection and evaluation of organ involvement in systemic amyloidosis.

**Methods:** We retrospectively included 83 participants, including 38 AL patients, 8 hATTR polyneuropathy patients, 28 PCD patients and 9 healthy controls. Whole-body PET/CT using [^18^F]FBP was performed in all participants, [^18^F]FDG PET was performed in 37 patients with AL and PCD, and the results were analyzed by visual and quantitative assessment. Biochemical, serum, and urine assays and histological analysis of tissue biopsies were performed.

**Results:** [^18^F]FBP SUV and TBR analysis showed comparable uptake in AL and hATTR-PN patients (p.A117S, p.V50M, p.K55N, p.T69AM, or p.H76R mutation carriers) and greater uptake than in PCD patients and control patients. Different regional [^18^F]FBP and [^18^F]FDG distributions were observed among the PCD, AL, and hATTR-PN groups. Both [^18^F]FBP and [^18^F]FDG enabled the detection of amyloidosis in patients with PCD before clinical detection of AL. [^18^F]FBP SUV and visual analysis provide comparable measures of organ involvement and were comparable to [^18^F]FDG and clinical assessment.

**Conclusions:** [^18^F]FBP PET detected organ amyloidosis in PCD, AL and hATTR-PN patients with high sensitivity and specificity and was more sensitive than [^18^F]FDG. Visual analysis and SUV analysis of [^18^F]FBP PET data provide comparable methods for evaluating organ involvement and are useful for noninvasively assisting in the early and accurate detection of systemic amyloidosis.

## Introduction

Systemic amyloidosis is a heterogeneous group of diseases characterized by localized or systemic deposition of insoluble extracellular fibrillary proteins in organs [1]. Systemic amyloidosis is categorized by precursor protein, and the most common are immunoglobulin light-chain amyloidosis (AL) produced by plasma cell dyscrasias (PCD) and transthyretin (TTR) protein produced predominantly in the liver (ATTR amyloidosis) [2]. PCDs, including monoclonal gammopathy (MGUS), multiple myeloma (MM), and smoldering MM, may develop AL. ATTR amyloidosis frequently results from age-related misfolding of wild-type TTR and, less commonly, from misfolding of a variant TTR from an autosomal dominant mutation of the TTR gene[3]. The symptoms of systemic amyloidosis are sometimes not specific and contribute to poor disease prognosis [1]. The survival of patients with AL amyloidosis is very short, ranging from 5 months to 2-3 years. Thus, it is extremely critical for the detection of amyloidosis at the earliest stage when therapy is still effective before severe organ damage occurs.

The gold standard of amyloidosis is biopsy histology. Noninvasive imaging approaches have facilitated the characterization of systemic amyloidosis in the early stages to guide intervention [4]. Scintigraphy using the calcification detecting tracer [^99m^Tc]3,3-diphosphono-1,2-propanodicarboxylic acid (DPD), [^99m^Tc]pyrophosphate, [^99m^Tc]HMDP [5, 6] and the [^123^I]serum amyloid P component [7] has been used to assist in the diagnosis of systemic amyloidosis. In addition, cardiac magnetic resonance has been valuable for detecting patients with cardiac amyloidosis. Positron emission tomography (PET) using [^18^F]NaF [8], the fibroblast activation protein inhibitor (FAPI) tracer evuzamitide ([^124^I]p5+14)[9], [^68^Ga]pentixa [10]), [^64^Cu]fibrin binding protein, [^18^F]fluorodeoxyglucose (FDG) and amyloid imaging tracers has been used in the clinical evaluation of systemic amyloidosis. [^18^F]FDG, which is based on the metabolic reprogramming of cells to favor glucose utilization within the glycolytic pathway and assesses inflammation by visualizing inflammation at sites with high glucose utilization in the brain and periphery, is commonly used in oncology practice. An earlier study reported the diagnostic and prognostic utility of [^18^F]FDG in patients with AL amyloidosis, localized amyloidosis [11-14], and pancreatic AL amyloidosis associated with MM [15, 16]. PET tracers such as [^18^F]flutemetamol, [^18^F]florbetapir (FBP) [17], [^18^F]florbetaben [18-20], and [^11^C]PIB [21], which are widely used for detecting amyloid-beta plaques in the brains of patients with Alzheimer’s disease [22], have emerged as valuable tools for the diagnosis of systemic amyloidosis. These tracers have shown prognostic utility in detecting cardiac amyloidosis (CA) in patients with hereditary ATTR (hATTR) [23-26] and AL [27, 28] and in identifying pulmonary involvement in individuals with AL [29-31]. [^18^F]FBP PET has been reported for diagnosis and follow-up in AL patients with cardiac or multiorgan involvement[17, 29, 30, 32-34] and ATTR patients[35]. [^11^C]PIB PET/CT has been used for treatment monitoring with tafamidis in patients with ATTR [36]. Moreover, [^18^F]florbetaben PET/CT is in a clinical trial [FLORAMICAR2] for differential diagnosis among AL-CA, ATTR, and mimicking conditions.

Thus far, a head-to-head comparative study of the utility of amyloid tracers and ^18^F]FDG PET for the early detection of amyloidosis in patients with PCD and AL has not been reported. The primary aim of this study was to evaluate and compare [^18^F]FBP and [^18^F]FDG for identifying organ involvement in AL and PCD and to determine which analysis is most suitable. Moreover, we included [^18^F]FBP imaging in hATTR-PN patients carrying p.H69R, p.A117S, p.K55N, or p.T69A mutations, for which no amyloid PET has been reported, as well as in the more common p.V50M mutation carrier.

## Methods

### Patient information

Eighty-three participants were included in this study, including 9 normal controls (NCs), 38 patients with AL, 8 patients with systemic hereditary transthyretin amyloidosis with polyneuropathy (hATTR-PN), and 28 patients with plasma cell dyscrasias-related PCD (including MGUS, MM, polyneuropathy, organomegaly, endocrinopathy, monoclonal protein, and skin changes [POEMS]). The participants were recruited from Huashan Hospital in Shanghai from 2022 to 2024 (detailed information is provided in **Tables 1** and **2**, **Table 1**). The participants volunteered to join the study cohort through an advertisement on a public board. The clinical workflow is shown in **SFig. 1**. Further cardiac ultrasound amyloid screening and bone marrow (BM) examinations were performed. If the results of the patient’s serum test were abnormal and clinical manifestations of suspected systemic amyloidosis, such as heart failure, elevated proBNP and cTnT, low voltage on the electrocardiogram, proteinuria, elevated ACR and PCR, renal impairment, liver damage, nerve damage, especially axonal damage, skin manifestations, tongue hypertrophy, or muscle damage, were detected, all 83 participants underwent [^18^F]FBP PET/computed tomography (CT). Among these patients, thirty-seven (21 with AL and 16 with PCD) underwent [^18^F]FDG PET/CT scans. The patients with hATTR-PN and normal controls did not undergo [^18^F]FDG PET.

**Table 1.** Demographic and biochemical marker information.

|  | NC | PCD | AL | hATTR |
| --- | --- | --- | --- | --- |
| n | 9 | 28 | 38 | 8 |
| [<sup>18</sup>F]FBP (n) | 9 | 28 | 38 | 8 |
| [<sup>18</sup>F]FDG (n) | 1 | 16 | 21 | 0 |
| Age | 58.9±7.9 | 61.0±12.4 | 61.5±10.6 | 55.4±13.7 |
| Sex (F/M) | 3/6 | 7/21 | 17/22 | 2/5 |
| AST (U/L) 9-50 |  | 21.2±18.7 | 26.0±19.4 | 20.1±4.5 |
| ALT (U/L) 15-40 |  | 21.8±24.8 | 29.6±28.9 | 22.8±2.7 |
| γ-GT (U/L) 10-60 |  | 44.7±52.0 | 47.7±33.0 | 18.0±5.2 |
| ALP (U/L) 45-125 |  | 91.1±48.8 | 93.3±66.6 | 65.0±14.8 |
| Total bilirubin (ng/ml) <26 |  | 7.3±3.6 | 12.3±8.0 | 7.4±1.8 |
| NT-proBNP (pg/mL) <219.3 | / | 1438.8±4749.0*** | 2938.1±6331.1 | 603.6±1045.5*** |
| sFLC κ (mg/L) 6.7-22.4 |  | 167.1±373.9 | 140.1±333.8 | 17.8±2.8 |
| sFLC λ (mg/L) 8.3-27 |  | 180.7±299.5 | 814.1±3388.1 | 20.9±4.9 |
| sFLC κ/λ 0.31-1.56 |  | 7.7±25.9 | 9.4±27.7 | 0.9±0.2 |

AL, light chain amyloidosis; AST, alanine aminotransferase; ALT, aspartate aminotransferase; ALP, alkaline phosphatase; F, female; γ-GT, gamma-glutamyltransferase; hATTR, hereditary transthyretin amyloidosis; M, male; NC, normal control; PCD, plasma cell dyscrasias; **sFLC**, serum free light chain; \*\*\*, p <0.0001

**Table 2.**
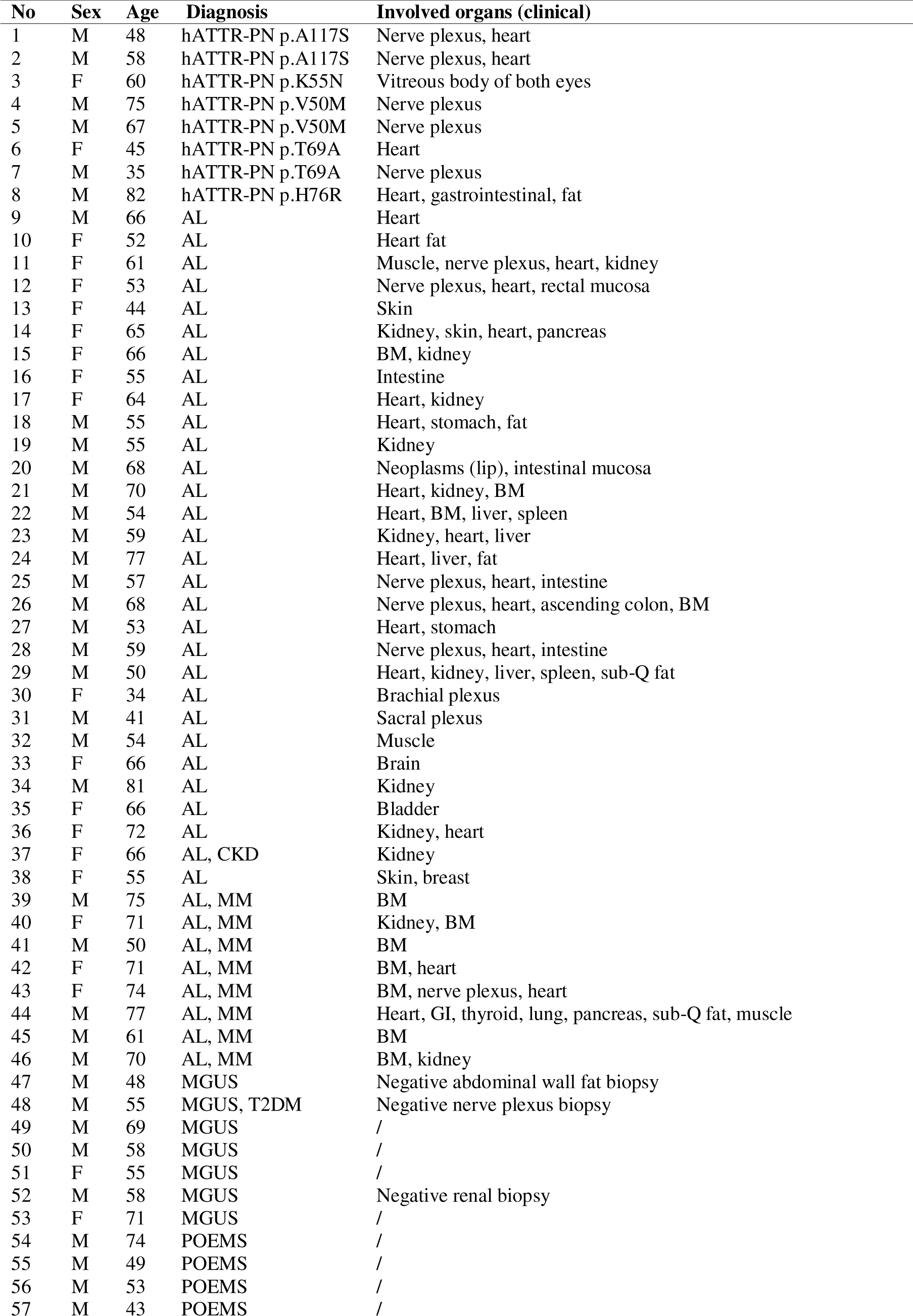
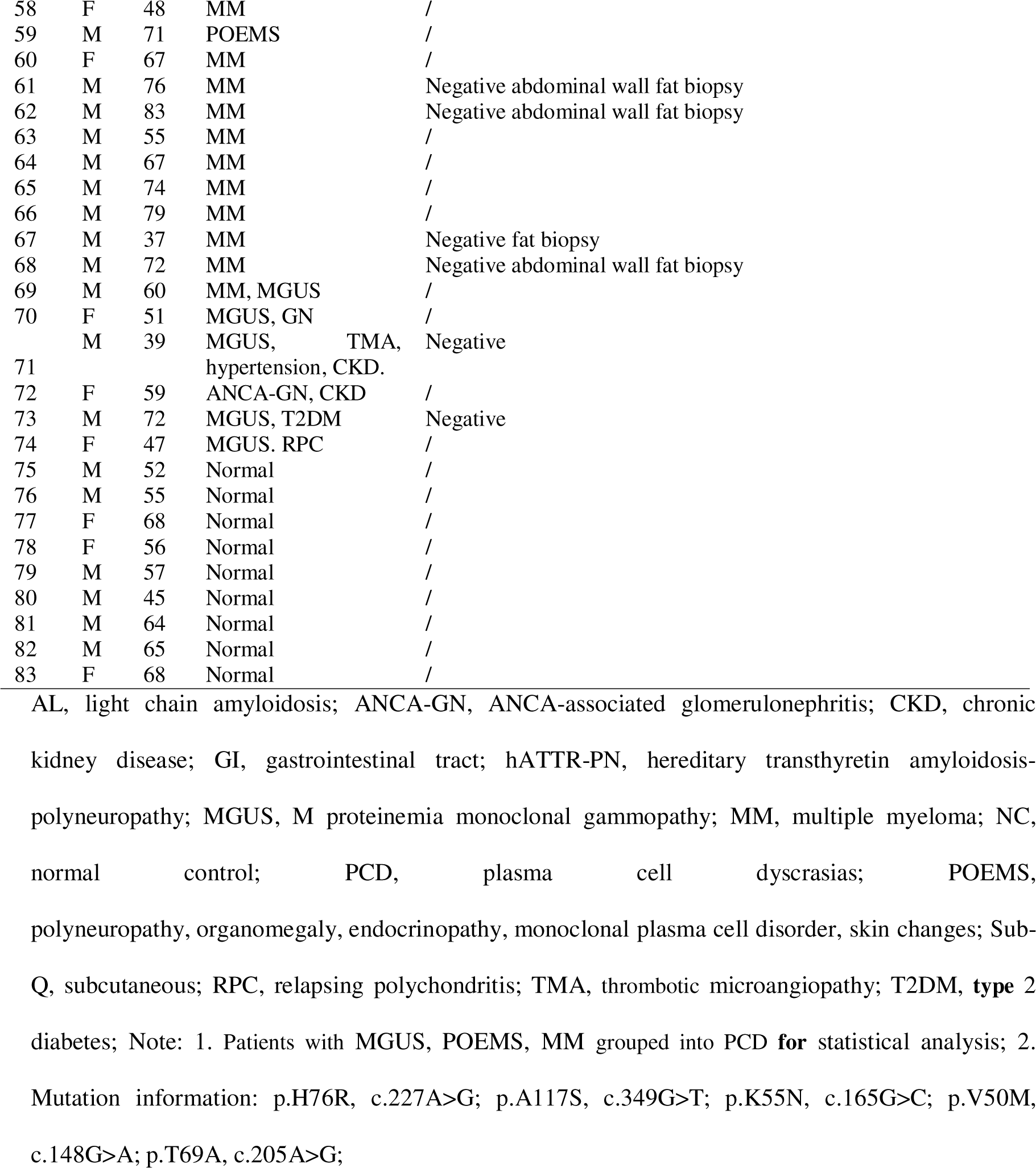
Characteristics of hATTR patients.

### Radiosynthesis

The [^18^F]FBP and [^18^F]FDG tracers were prepared in the Department of Nuclear Medicine, PET Center, Huashan Hospital, Fudan University, as described earlier [37-39]. [^18^F]FBP (0.56 GBq/ml) was radiosynthesized from its precursor via a fully automated procedure [37-40]. [^18^F]FDG (1.48 GBq/ml) was prepared in the radiochemistry facility under Good Manufacturing Practices requirements [37, 38].

### PET/CT

[^18^F]FBP and [^18^F]FDG PET/CT were conducted (Biograph Truepoint HD 64 PET/CT or Biograph mCT Flow, Siemens, Germany) with previously described parameters [39]. For [^18^F]FBP PET, subjects (n=83) were intravenously injected with 0.37-0.55 MBq/kg [^18^F]FBP. At 50 min after the injection, a 2 min static PET acquisition for the body in 3D+time of flight (TOF) mode with a 12.18 s CT scan (120 kV, 150 Ma) for attenuation correction was performed [41]. After the scan, a 10 min static positron emission tomography (PET) scan of the brain was performed in 3D+TOF mode with a 12.18 s CT scan (120 kV, 150 Ma) for attenuation correction. For [^18^F]FDG PET, 21 AL and 16 PCD patients were intravenously injected with 0.37-0.55 MBq/kg [^18^F]FDG. At 90 min after the injection, a 2 min PET acquisition scan of the body in 3D+TOF mode with a 12.18 s CT scan (120 kV, 150 Ma) for attenuation correction was performed. After the scan, a 10 min PET acquisition scan for the brain was performed in 3D+TOF mode with a 12.18 s CT scan (120 kV, 150 Ma) for attenuation correction. After the acquisition, all brain PET images were reconstructed utilizing a backprojection + TOF algorithm with the following parameters: image size= 256×256, Gaussian filter, zoom factor 2, and full width at half maximum (FWHM) = 3.5 [39]. For whole-body PET, images were reconstructed using a Gaussian filter, full width at half maximum (FWHM) = 4, zoom = 1, image size = 200×200, recon method Trux+TOF, iteration = 4.

### Image analysis

Two experienced PET specialists used the binary visual reading method to analyze the involved organs on [^18^F]FBP and [^18^F]FDG PET images. When the evaluations of the two PET specialists were not consistent, a senior PET specialist provided further evaluation [42]. The positivity of the PET scans in both the brain and the periphery was visually analyzed. Standard uptake value (SUV) and target-to-background ratio (TBR) analyses of [^18^F]FBP and [^18^F]FDG PET data were performed for 20 organs, including the parotid gland, tongue, submandibular gland, thyroid, humeral head, humerus, muscle, lymph node, lung, heart, mediastinal blood pool (MBP), liver, stomach, spleen, pancreas, kidney, intestine, fat, and BM. Bladders were not included in the analysis because urine accumulation confounded the results. Since only one patient showed positive uptake in the spinal cord, retina, breast and skin, we did not perform an analysis of these organs. The TBR was computed using the MBP (circle, diameter 1 cm, **SFig. 3**) as the reference region for both [^18^F]FBP and [^18^F]FDG PET.

### Biochemical analysis and staining of biopsy tissues

Biochemical analysis (serum, urine) was performed for different organs. In the liver, alanine transaminase (ALT), aspartate transaminase (AST), gamma-glutamyl transferase (r-GT), alkaline phosphatase (ALP), and total bilirubin were measured following standard procedures. For the heart, cardiac troponin T (cTnT), N-terminal pro-B-type natriuretic peptide (NT-proBNP), heart-type fatty acid-binding protein (hFABP), and serum free light chain (SFLC) were measured following standard procedures. For the kidney, the estimated glomerular filtration rate (eGFR) was measured following standard procedures. For urine, the urine protein, kappa and lambda light chain, as well as the Bence-Jones protein, were measured following standard procedures.

Genetic testing was performed for participants with ATTR to confirm mutations (xymedlab, Genery Biotechnology, Amcarelab, and Kangso Medical, China). A blind test of abdominal wall fat biopsy or rectal biopsy was performed if not available. Biopsies were collected from the retina, muscle, liver, fat tissue, skin, liver, kidney, bladder, lung and brachial plexus. The biopsy site was determined based on the imaging and clinical examination results. The site of amyloid deposition was used if available. Congo red staining and hematoxylin and eosin (H&E) staining were performed on the paraffin-embedded fixed tissue biopsy following standard procedures. Kappa and lambda immunofluorescence staining were performed on frozen tissue biopsy sections following standard procedures. The stained sections were scanned at ×20, ×40 and ×100 magnification using a Nikon anti-mould microscope (551 DS-F1, Nikon Instruments). Images were adjusted using ImageJ (NIH).

### Statistics

Statistical analysis was performed using GraphPad Prism 10 (GraphPad). The nonparametric MannLWhitney test was used for comparisons between two groups. Two-way ANOVA was performed for values from different regions of different groups. Correlations between different readouts were analyzed by using nonparametric Spearman’s rank analysis. The values are expressed as the mean ± standard deviation. The significance level was set at p<0.05.

## Results

### Demographic characteristics and clinical assessments

The demographic information of the patients and normal controls is listed in **Table 1**. In total, 83 participants, including 9 NC (58.9±7.9), 38 AL (61.5±10.6), 28 PCD (61.0±12.4), and 8 ATTR (55.4±13.7) participants, were included in the retrospective study. The clinical workflow is shown in **SFig. 1**. Detailed information on the diagnosis and regions involved in amyloidosis based on clinical diagnosis is listed in **Tables 1** and **2**. No difference in age was found between the groups. Among the PCD patients, 11 had MM, 13 had MGUS (with other complications), and 4 had POEMS. Among the 8 patients with hATTR-PN, 1 patient carrying p.H76R (c.227A>G), 2 patients carrying p.A117S (c.394G>T), 2 patients carrying p.V50M (c.148G>A), 2 patients carrying p.T69A (c.205A>G), and 1 patient carrying p.K55N (c.165G>C) were included. The clinical symptoms reported in the literature are consistent with these cases. One hATTR-PN (p.H76R) patient clinically manifested with first-degree atrioventricular block, cardiac radiofrequency ablation, prostatic hyperplasia and pleural effusion. The two hATTR-PN (p.A117S) patients clinically manifested multiple sensorimotor peripheral neuropathies, with amyloidosis in the peripheral nerve plexus and heart and distal cold limbs. Two hATTR-PN (p.V50M) patients exhibited amyloidosis in the peripheral nerve plexus and sensorimotor peripheral impairment. One patient with p.T69A had amyloidosis in the peripheral nerve plexus with normal cardiac function (45 y, female), while the other two had cardiac amyloidosis in the gastrointestinal system and sensorimotor peripheral impairment (58 y, male). The hATTR-PN (p.K55N) patient clinically manifested with vitreous amyloidosis in both eyes. The levels of biochemical markers in the PCD, AL and hATTR-PN groups are shown in **Table 1** and **STable 1**. The AST, r-GT, ALP, total bilirubin, hFABP, SFLC, creatinine, and eGFR levels were comparable between the groups (**Table 1** and **STable 1**). Significantly greater levels of NT-proBNP were detected in the AL group than in the PCD and hATTR-PN groups (**Table 1**, p<0.0001, p<0.0001).

### [^18^F]FBP detected organ amyloidosis in PCD, AL and hATTR-PN patients with high sensitivity and specificity

We first assessed the distribution pattern and compared the SUV and TBR of [^18^F]FBP between the NC group and patients with AL, PCD and hATTR-PN. The TBR was quantified using MBP as a reference (**SFig. 3**). The highest [^18^F]FBP uptake was observed in the liver, followed by the BM, in PCD, AL, and hATTR-PN patients (p.A117S, p.V50M, p.T69A, or p.K55N p.H76R, mutation) (**Figs. 1-2**). All the AL and hATTR-PN patients (p.A117S, p.V50M, p.T69A, or p.K55N p.H76R, mutations) were positive according to [^18^F]FBP PET, although different patterns and distributions of [^18^F]FBP uptake were found in the heart, kidney, BM, fat, and muscle of patients with hATTR-PN carrying the p.A117S mutation (**Fig. 1a-c**). We observed positive [^18^F]FBP uptake in the BM, stomach, and fat in the pelvic floor, kidney, and muscle of patients with hATTR-PN carrying the p.T69A mutation (**Fig. 1d-g**). Patients with hATTR-PN carrying the p.K55N mutation showed positive [^18^F]FBP uptake in the vitreous body of both eyes, in addition to uptake in the heart, muscle, lung and BM (**Fig. 1h-k**) and in the kidney (l) and muscle (m) of hATTR-PN patients carrying the p.V50M mutation (age 67 y, male, **Fig. 1l-n**). Positive [^18^F]FBP uptake was detected in the heart, tongue, muscle, kidney, BM and fat of the hATTR-PN patient harboring the p.H76R mutation (**Fig. 1o-u**).

**Fig 1.**
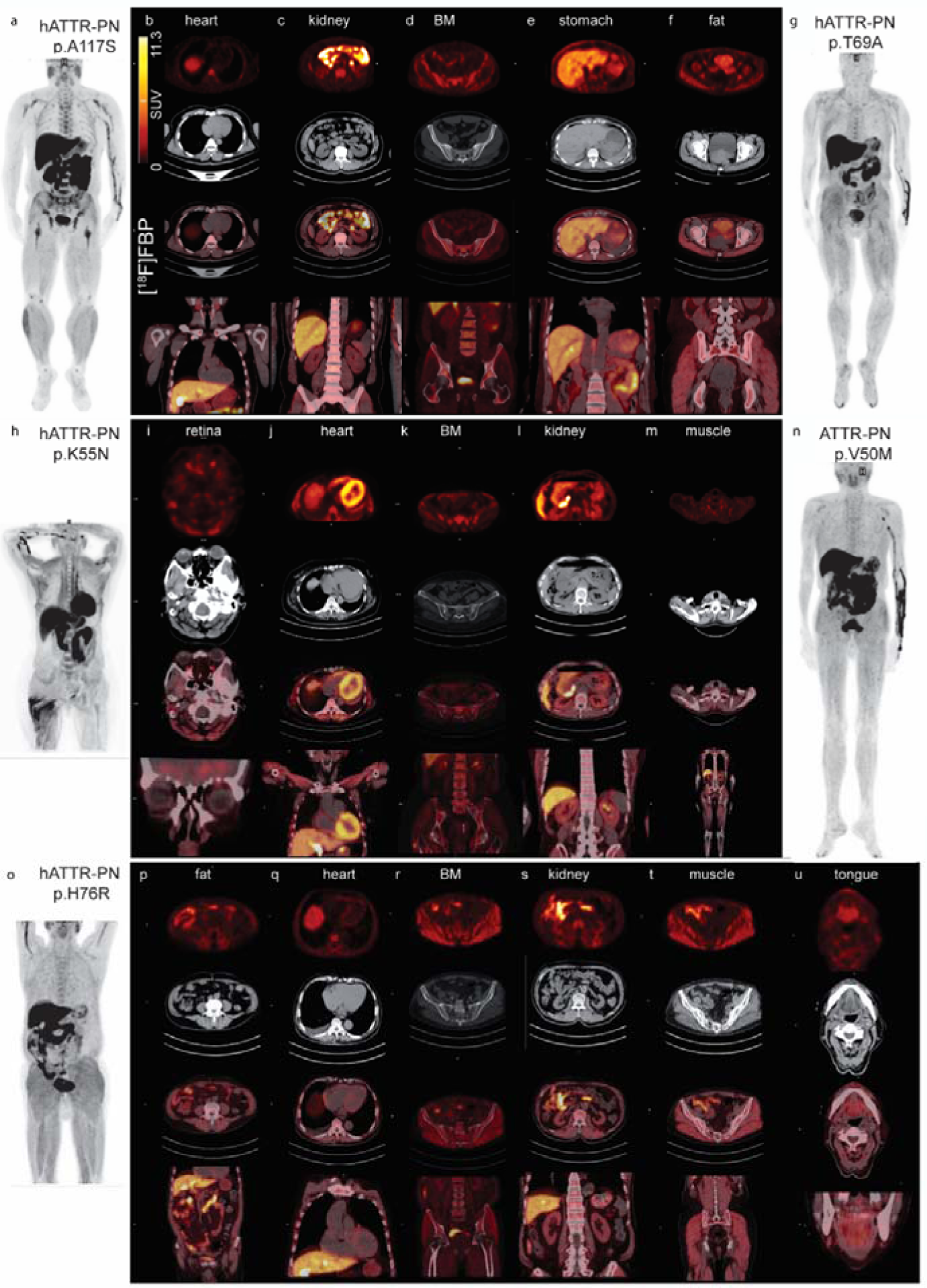
[^18^F]FBP imaging in hATTR-PN patients. (a, g, h, n, o) Maximum intensity projection (MIP) of [^18^F]FBP whole-body scans of hATTR-PN patients. Transaxial PET, CT, transaxial and coronal images of overlay images of the heart (b), kidney (c) and bone marrow (d) from a patient with hATTR-PN (p.A117S, aged 48 y, male, a); of the stomach (e), and fat in the pelvic floor (f) from a patient with hATTR-PN (p.T69A, aged 45 y, female, g); of the retina (i), heart (j) and BM (k) from a patient with hATTR-PN (p.K55aN, aged 60 y, female, h); and of the kidney (l) and muscle (m) from a hATTR-PN patient (p.V50M, aged 67 y, male, n). and fat (p), heart (q), kidney (r), muscle (s), and tongue (u) of a hATTR-PN patient (p.H76R, aged 82 y, male, o).

**Fig. 2.**
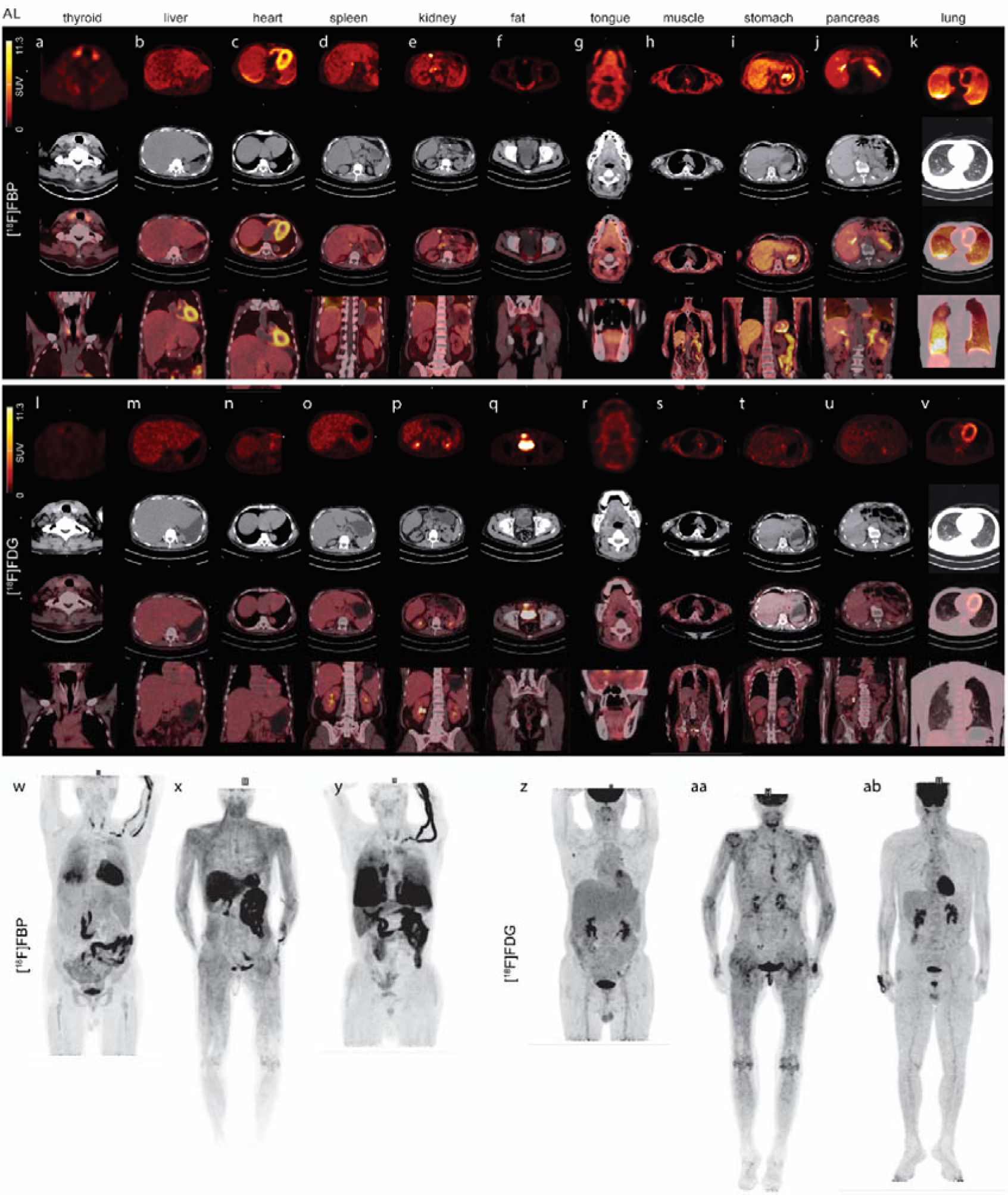
Head-to-head comparison of [^18^F]FBP and [^18^F]FDG PET in the same AL patients. (a-v) Representative transaxial view of [^18^F]FBP and [^18^F]FDG PET, CT, transaxial and coronal view of overlay images of the thyroid (a, l), heart (b, m), liver (c, n), spleen (d, o), kidney (e, p), and fat in the pelvic floor (f, q), in one AL patient (aged 77 y, male, w,z); tongue (g, r), muscle (h, s) of one AL patient, stomach (i, t), pancreas (j, u), and lung (k, v) of one AL patient (x, aa); (w-y) Maximum intensity projection (MIP) of [^18^F]FBP (z-ab) and [^18^F]FDG whole-body scans of the patients.

In AL patients, positive [^18^F]FBP uptake was detected via both visual and SUV analysis in the tongue, thyroid, heart, liver, spleen, kidney, subcutaneous (Sub-Q) fat, and fat in the pelvic floor, muscle, stomach, breast, skin, pancreas and lung (**Fig. 2**). A few AL patients showed positive [^18^F]FBP uptake in the central nervous system, which might be due to deposition of cerebral amyloid angiopathy or amyloid-beta (**SFig. 2**). In addition, [^18^F]FBP enabled the detection of amyloidosis in patients with PCD (including patients with MGUS, POEMS, and MM) at an early stage before the clinical diagnosis of amyloidosis (**Fig. 3)**. Positive [^18^F]FBP uptake was detected in the kidney, BM, humerus, fat tissue, heart, thyroid, stomach, spleen, and lymph nodes of patients with PCD by both visual and SUV analysis. A different pattern of % organ involvement by [^18^F]FBP was observed in different groups: highest in muscle, BM and kidney (85%+) of hATTR-PN, kidney of AL (60%+), and BM of PCD (85%+) (**Fig. 5g-i, SFig. 5a**).

**Fig 3.**
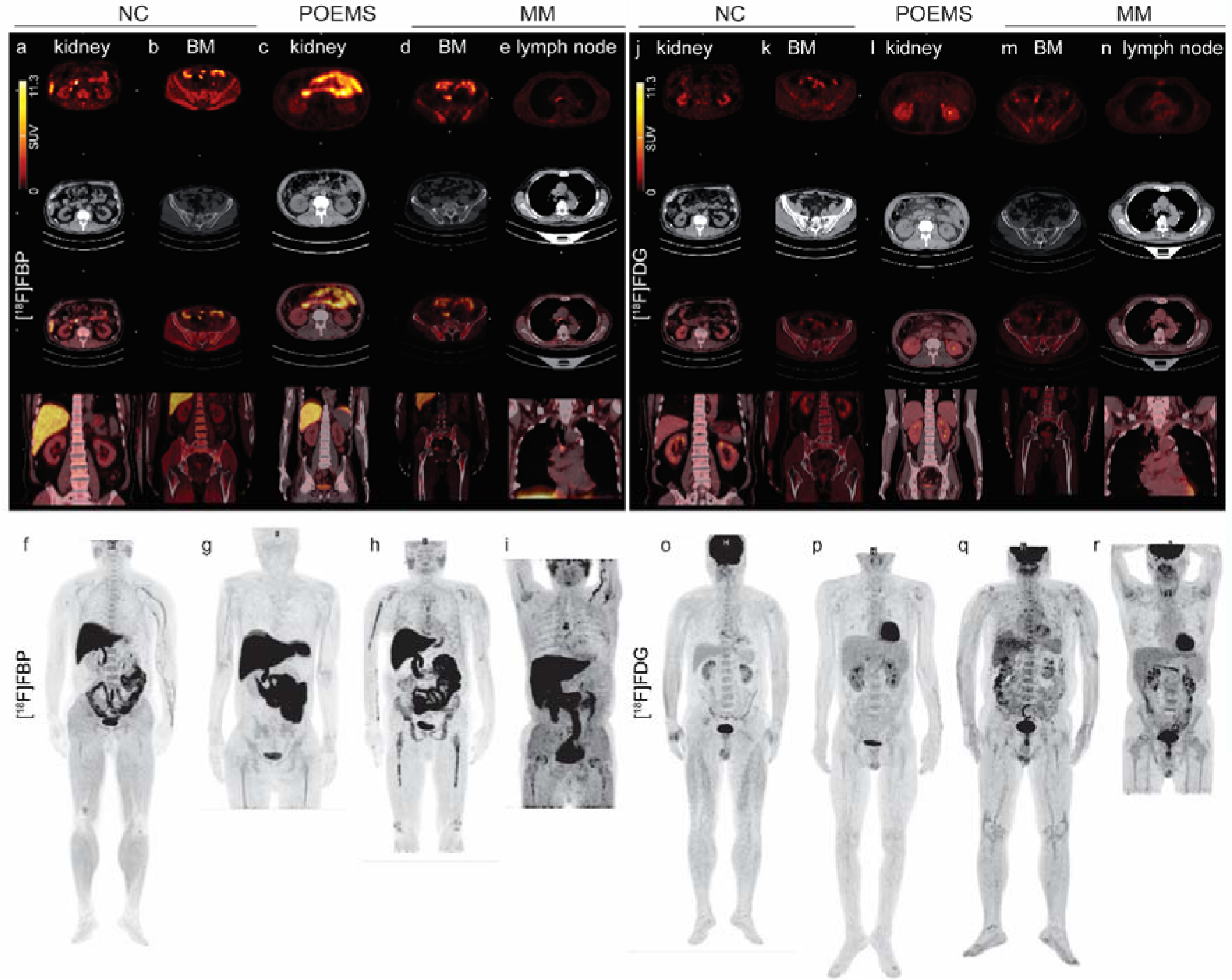
[^18^F]FBP and [^18^F]FDG detection in amyloidosis patients with plasma cell dyscrasias (POEMS and MM). (a-e) Transaxial view of [^18^F]FBP and [^18^F]FDG PET, CT, transaxial and coronal view of overlay images in the kidney (a, j) and bone marrow (BM, b, k) in the NC (control, f, o); kidney (c, l) of one POEMS patient (g, p); BM (d, m) of one MM patient (h, q); lymph node (e, n) of one MM patient (i, r); and maximum intensity projection (MIP) of [^18^F]FBP (f-i) and [^18^F]FDG (o-r) whole-body scans of NC, POEMS and MM patients. MM, multiple myeloma; POEMS, polyneuropathy, organomegaly, endocrinopathy, monoclonal protein, skin changes.

SUV analysis revealed greater regional [^18^F]FBP uptake in the heart, lung, and thyroid in the AL group than in the PCD group and in the heart than in the NC group (**Fig. 4**). TBR analysis revealed greater [^18^F]FBP uptake in the heart, lung, muscle, liver and thyroid in the AL group than in the PCD group; in the heart, liver, muscle, thyroid, fat, and BM in the AL group than in the NC group; and in the fat and heart in the AL group than in the hATTR group (**Fig. 4**). In addition, TBR analysis revealed greater [^18^F]FBP uptake in the liver in the hATTR group than in the NC group.

**Fig 4.**
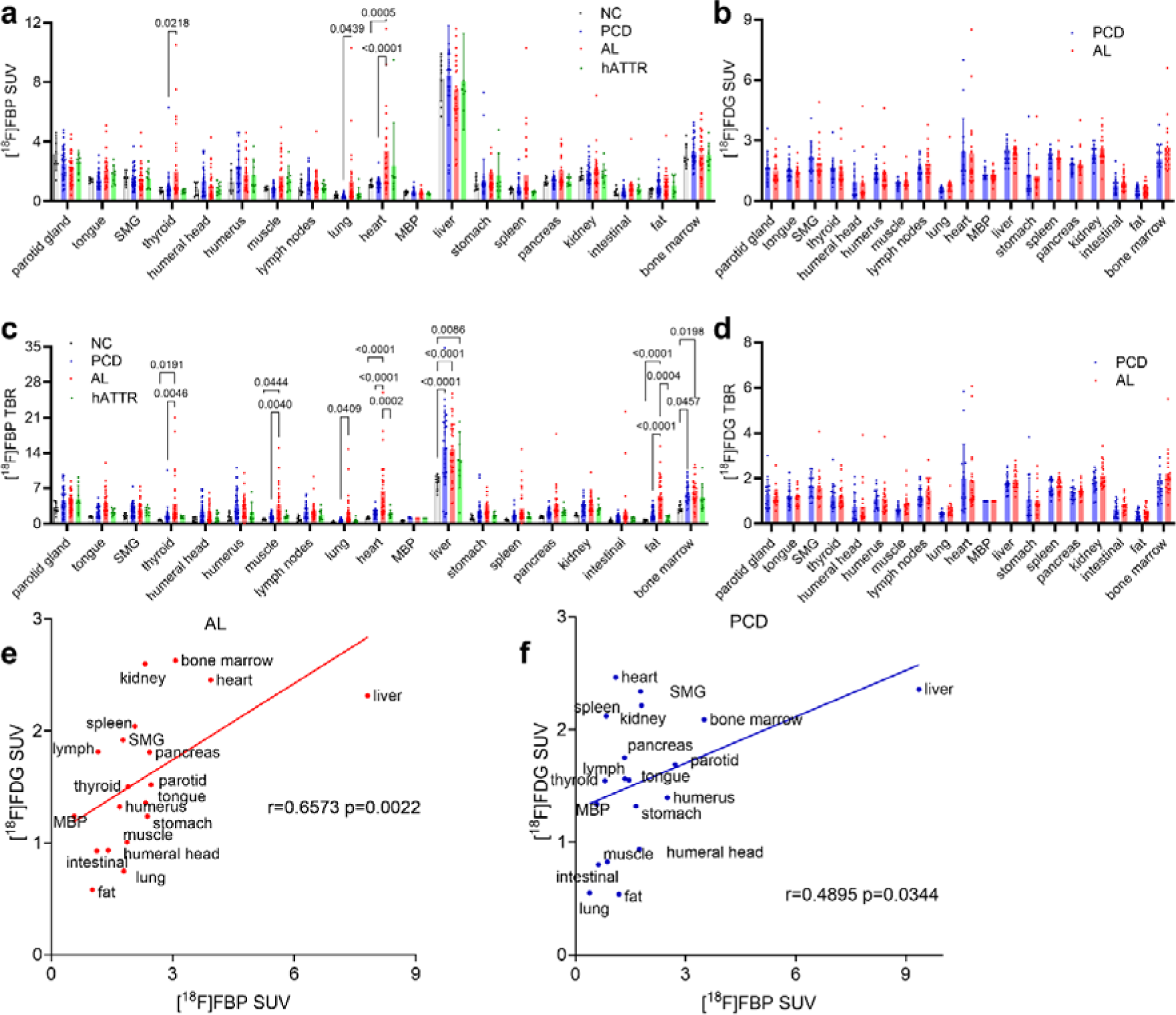
Regional uptake of [^18^F]FBP and [^18^F]FDG in the NC group. PCD, AL, and hATTR-PN patients. (a, b) SUV analysis of [^18^F]FBP and [^18^F]FDG; (c, d) TBR analysis of [^18^F]FBP and [^18^F]FDG; (e, f) correlation between regional [^18^F]FBP SUV and [^18^F]FDG SUV in the AL and PCD groups. PCD. plasma cell dyscrasias; mediastinal blood pool (MBP); submandibular gland (SMG).

### [^18^F]FDG regional distribution and comparison between SUV and visual and clinical assessments of organ involvement

[^18^F]FDG PET was performed for patients at risk of having a tumor or MM. Next, we assessed the distribution pattern of [^18^F]FDG and compared the SUV and TBR in patients with AL or PCD (not in NC or hATTR-PN patients). The highest [^18^F]FDG uptake was observed in the liver, followed by the BM, in the PCD and AL groups (**Figs. 1-3**). All the AL patients had positive [^18^F]FDG PET results according to visual, SUV, and TBR analyses. In addition, [^18^F]FDG SUV analysis enabled the detection of amyloidosis in patients with PCD (including patients with MGUS, POEMS, and MM) at an early stage before the clinical diagnosis of amyloidosis. The % organ involvement was greatest in the heart (50%+), kidney (45%+) and BM (35%+) of the ALs and in the BM (65%+) and humerus (45%+) of the PCD patients (**Fig. 5g, h, SFig 5b**). SUV and TBR analyses revealed that the regional uptake of [^18^F]FDG was comparable between the AL group and the PCD group (**Fig. 4b, d**).

**Fig. 5.**
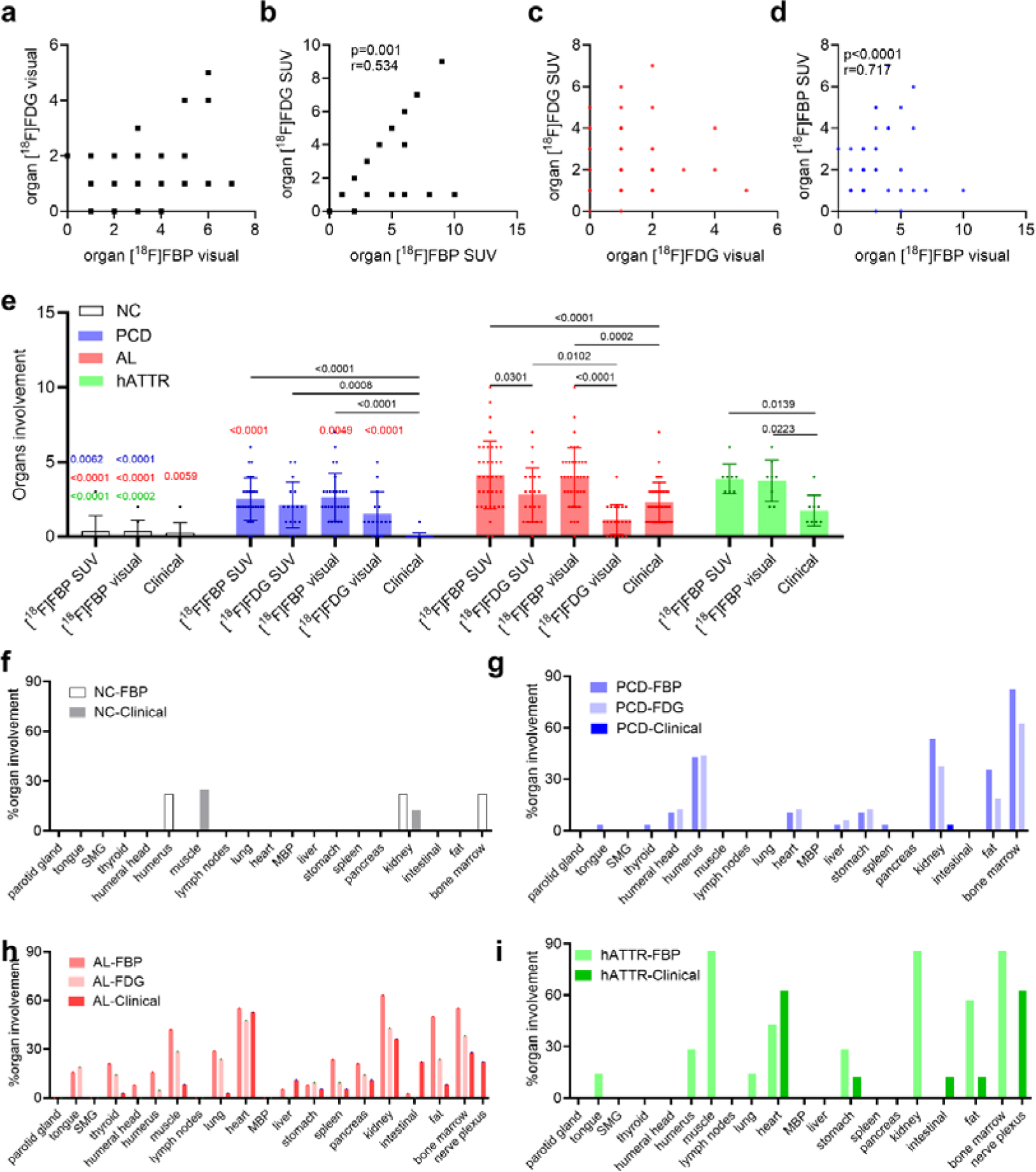
Association between clinical assessment and visual and SUV analysis of [^18^F]FBP and [^18^F]FDG on organ involvement. In 21 AL and 16 PCD patients, (a) [^18^F]FBP visual and [^18^F]FDG visual; (b) [^18^F]FBP SUV and [^18^F]FDG SUV; (c) [^18^F]FDG visual and SUV; and (d) [^18^F]FBP visual and SUV. (e) Comparison of the SUV and visual analysis of [^18^F]FBP, [^18^F]FDG PET and clinical diagnosis in NC, PCD, AL and hATTR-PN patients; p values in black comparison with the group; p values in red vs. AL; blue vs. PA; green vs. hATTR-PN. (f-i) % organ involvement according to SUV analysis of [^18^F]FBP, [^18^F]FDG and clinical assessment in the NC group. PCD, AL, hATTR-PN patients; PCD. plasma cell dyscrasias; mediastinal blood pool (MBP); submandibular gland (SMG).

### Head-to-head comparison between [^18^F]FBP and [^18^F]FDG and clinical diagnosis

SUV, TBR and visual analysis of [^18^F]FBP all showed high sensitivity (1) and specificity (0.97) in detecting amyloidosis in organs (**Fig 4**). In comparison with SUV analysis, TBR analysis of [^18^F]FDG, but not visual analysis, showed high sensitivity (63%) and specificity in detecting amyloidosis (**Fig 4**). The number of organs involved according to the SUV and visual analysis of [^18^F]FBP in the AL group was greater than that in the PCD group (p < 0.0001, p = 0.0049) and NC group (p < 0.0001, < 0.0001) and comparable to that in the hATTR-PN group (**Fig. 5e, STable 3**). The number of organs involved according to the SUV and visual analysis of [^18^F]FBP in the PCD patients was greater than that in the NCs (p = 0.0002, < 0.0001) (**Fig. 5e, STable 3**). The number of organs involved according to the SUV and visual analysis of [^18^F]FBP in the hATTR-PN group was greater than that in the NC group (p < 0.0001, p = 0.0002) (**Fig. 5e, STable 3**).

In comparison, the visual analysis of [^18^F]FDG was not different from that of clinically detected organs. A greater number of involved organs were detected by SUV analysis of [^18^F]FDG than by visual analysis and clinical assessment in the AL group (p = 0.0102, p = 0.0002; **Fig. 5e**; Table S3). A greater number of involved organs were detected by SUV analysis of [^18^F]FDG than by clinical assessment in the PCD group (p = 0.0008). The SUV and visual analysis of [^18^F]FDG were significantly lower than those of [^18^F]FBP in the AL group (p = 0.0301, p < 0.0001; **Fig. 5e; STable. 3**).

SUV analysis revealed that [^18^F]FDG and [^18^F]FBP were positively correlated with the number of organs involved in AL and PCD patients (p=0.001, r=0.534, Spearman rank analysis, **Fig 5a, b**), but this correlation was not detected by visual analysis. Regional SUV analysis revealed that [^18^F]FDG and [^18^F]FBP were correlated in both AL (p=0.0022, r=0.6573) and PCD patients (p=0.0344, r=0.4895). The [^18^F]FDG SUV and [^18^F]FBP SUV were correlated within the humeral head, humerus, muscle, lung, stomach, spleen, pancreas, intestine, and fat (**STable. 4**). In addition, the SUV and visual analysis of [^18^F]FDG did not correlate, while the SUV and visual analysis of [^18^F]FBP were positively correlated with the number of organs involved in AL and PCD patients (p=0.0001, r=0.717; Spearman rank analysis, **Fig. 5c, d**).

### Staining of biopsy tissue sections validated the presence of amyloidosis

Pathological examination was performed on all the AL and hATTR-PN patients and a proportion of the PCD patients. Fat tissue is the most commonly used tissue for biopsy. Amyloid deposits were identified as amorphous, waxy-appearing, pink material on hematoxylin and eosin (H&E) staining with 1) apple-green birefringence by polarized light microscopy with Congo red staining and 2) a pink-to-red color under a transmitted-light microscope with Congo red staining (**Fig. 6a-ab**). For the fixed liver biopsy tissue of the AL patient (age 77 y, male, PET data shown in **Fig. 1b**), the large amount of eosin-structured material deposits in the hepatic sinusoids of a small amount of punctured tissue was consistent with amyloidosis (**Fig. 6a, b**). Fixed liver tissue from the same AL patient was stained positively with Congo red, and typical birefringence was visible under a polarized light microscope (**Fig. 6c, d**).

**Fig. 6.**
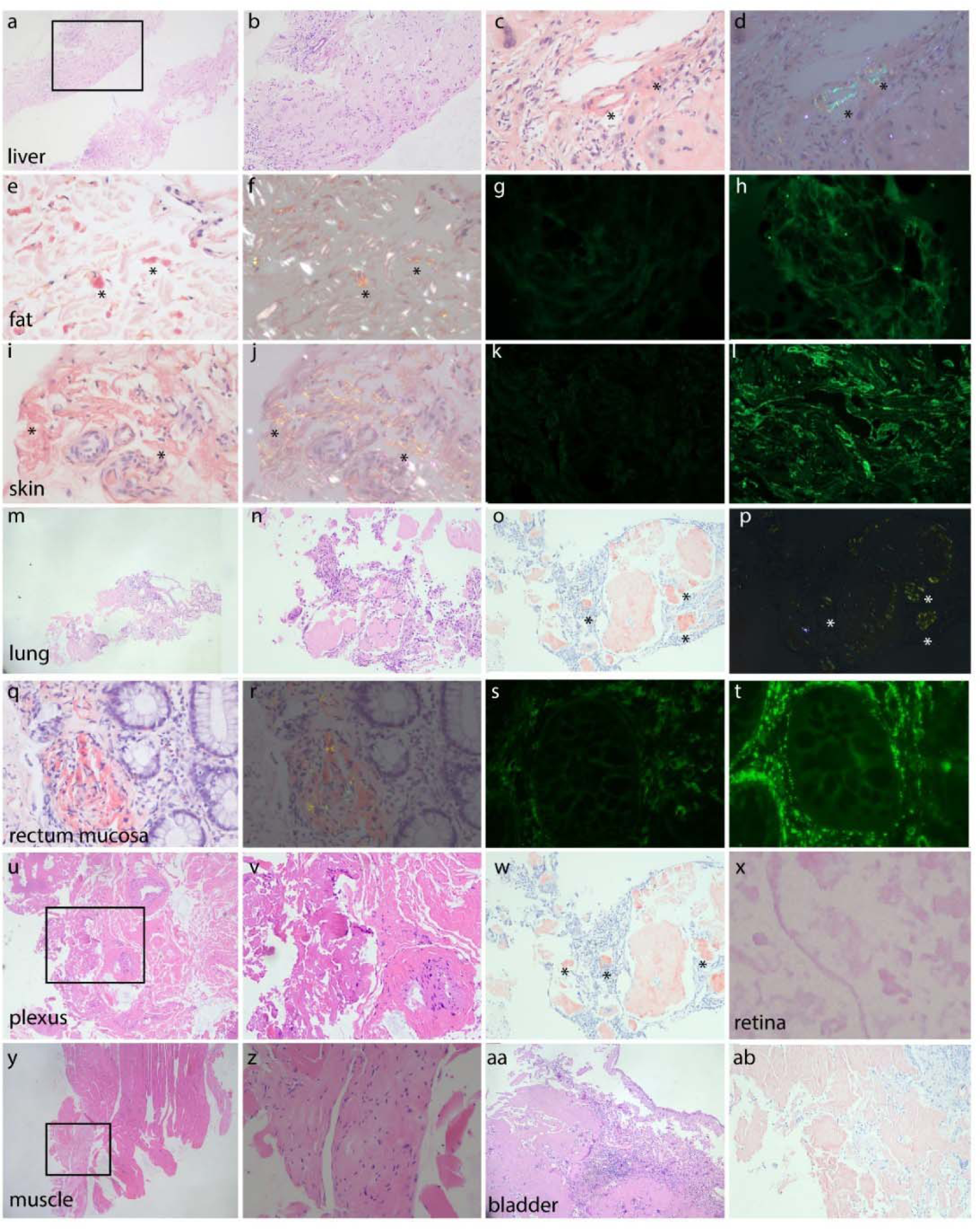
Staining of biopsy tissue from systemic amyloidosis patients. In the liver (a-d), fat (e-h), skin (i-l), lung (m-p), rectum mucosa (q-t), brachial plexus (u-w), retina (x), muscle (y, z), and ladder (aa, ab). Hematoxylin and eosin (H&E, a, b, m, n, u-v, x-aa) and Congo red (c-f, i-j, o-r, w, ab) staining demonstrating the reactivity of amyloid deposits with apple-green birefringence to polarized light. Kappa (g, k, s) and lambda (h, l, t). Panels b, v, and z are magnified views of a, u, and y, respectively.

Congo red staining was performed on the fixed Sub-Q fat biopsy tissue from patients with AL and revealed positive birefringence (**Fig. 6e, f**, example from one patient aged 55 y, female); immunofluorescence analysis of frozen Sub-Q fat biopsy tissue sections revealed a negative Kappa and positive Lambda staining pattern, indicating amyloidosis (**Fig. 6g, h**). Congo red staining of paraffin-embedded fixed skin biopsy tissue sections from AL patients revealed obvious birefringence under polarized light microscopy (**Fig. 6i, j**. aged 55 y, female), negative kappa staining, and positive lambda staining of frozen skin biopsy tissue sections, indicating amyloidosis (**Fig. 6k, l**). For patients with flaky erosions, submucosal hematomas, blood blisters, ecchymoses, nodular protrusions or irregular ulcers under colonoscopy, biopsy of the rectum mucosa was performed. Congo red staining of the paraffin-embedded fixed rectum mucosa biopsy section and positive Kappa staining and positive Lambda staining of the frozen rectum mucosa tissue section confirmed the presence of amyloid deposits (**Fig. 6q-t**, aged 68 y, male).

## Discussion

We demonstrated the utility of [^18^F]FBP in visualizing amyloidosis in AL and hATTR-PN patients (with p.A117S, p.V50M, p.K55N, p.T69A, or p.H76R mutations) as well as in the early detection of PCD. In addition, we showed that visual analysis and SUV analysis of [^18^F]FBP provided comparable identification of organ involvement in PCD, AL, and hATTR-PN patients. [^18^F]FBP PET showed greater sensitivity in detecting organ involvement in amyloidosis than [^18^F]FDG PET, especially visual analysis, in AL and PCD patients.

We detected increased [^18^F]FBP uptake in hATTR-PN patients (p.A117S, p.V50M, p.K55N, p.T69A, p.H76R) compared to that in normal controls. Earlier studies using dynamic [^18^F]FBP and [^18^F]florbetaben PET showed the possibility of differentiating patients with AL from those with ATTRwt [34], probably due to the difference in the affinity of the tracers for AL fibrils compared to that for TTR. In addition, another study showed that late static [^18^F]florbetaben PET scan at 50 minutes postinjection was able to discriminate cardiac involvement due to AL amyloidosis from ATTR amyloidosis and from other non-CA conditions mimicking infiltrative disorders [34]. In the current study ([^18^F]FBP PET scan at 50 minutes), the uptake in the organs of hATTR-PN patients was comparable to that of AL patients.

The most common genetic variant in hATTR is V30M (p. V50M) and T60A worldwide [43, 44]. Several PET studies have reported greater uptake in patients with hATTR-CA carrying V30M (p. V50M) compared to healthy controls by using [^18^F]FBP [45], [^18^F]flutemetamol [46, 47], and [^11^C]PIB [21, 23]. Cerebellum and cerebral amyloid deposition in long-term hATTR V30M (p.V50M) patients has been previously reported by using [^18^F]flutemetamol PET [47]. In our study, no CNS involvement was observed in hATTR-PN (p.V50M) or hATTR-PN harboring other mutations. To date, no PET images using amyloid tracers in hATTR-PN patients carrying p.A117S, p.K55N, p.H76R or p.T69A mutations have been reported in prior studies. Positive [^11^C]PIB uptake has been reported in various organs of hATTR patients carrying G47R, T60A, or D18G mutations [21]. The hATTR-PN p.A117S (A97S) mutation has been reported in a few studies in East Asian populations but has not been reported in Caucasians [48-51]. hATTR-PN p.T69A (T49A) patients were reported in two studies from France and Italy (Sicily region) [52, 53], and hATTR-PN p.H76R (H56R) patients were reported once with geographic kindreds in the USA [54] and were both reported for the first time in the East Asian population. Thus far, only one case report on an hATTR-PN p.K55N (K35N) patient with vitreous amyloidosis, whose clinical manifestations were similar to those in the present study, has been published [55].

Scintigraphy with [^99m^Tc]DPD and HMDP was found to be suboptimal in TTR-CA patients with V30M (p.V50M) or p.F64L mutations [56] but positive in, e.g., E89Q, A36P, T49A, F33V, and I68L mutation carriers [57, 58]. These findings indicated the unique value of PET using amyloid tracers such as [^18^F]FBP in detecting amyloid deposits in patients with hATTR. Differences in the binding sites of amyloid PET tracers on amyloid-beta fibrils have previously been described in sporadic and autosomal dominant Alzheimer’s disease [59, 60]. An ex vivo study showed that the amyloid tracer [³H]PIB and the flutemetamol analog cyano-flutemetamol were able to detect AL and ATTR amyloid deposits [61, 62]. Further study on the binding affinity of the amyloid tracer for ATTR fibrils with different mutations will be informative.

Our data on organ involvement detected by [^18^F]FBP were in line with previously reported findings of amyloid-positive organs in the AL and hATTR-PN [2, 34, 35, 63, 64] and were greater than the clinical findings. An earlier study showed cardiac uptake with [^18^F]FBP PET in AL patients with cardiac involvement [17, 30], multiorgan involvement in patients with AL [29, 32-34], and the diagnosis and follow-up of ATTRwt patients [35]. An earlier study reported the utility of [^18^F]florbetaben [20] and [^18^F]florapronol [65] PET-CT in detecting AL amyloidosis in patients with MM and Waldenström’s macroglobulinemia [66]. [^18^F]FDG PET/CT is commonly used for diagnostic and staging imaging in MGUS, MM and other types of PCD [15]. [^18^F]FDG was able to detect pancreatic AL amyloidosis [67] and secondary gastrointestinal amyloidosis [68] in patients with MM, in pulmonary amyloidosis [69] and CA [13] in patients with AL.

[^18^F]FBP revealed organ involvement via both visual and SUV analysis in the tongue, thyroid, heart, humerus, liver, spleen, kidney, sub-Q fat, and fat in the pelvic floor, muscle, stomach, intestine, breast, skin, pancreas, brain, spinal cord and lung. Moreover, different regional [^18^F]FBP distributions were observed among the AL, PCD, and hATTR-PN groups and were more widespread in the AL group. The highest % involvement was observed in the muscle (85%+), kidney (85%+), and BM (85%+) of hATTR-PNs; BM (80%+), kidney (60%+), and humerus (45%+) involvement in PCD patients; and kidney (65%+), BM (55%+) and heart (50%+) involvement in AL patients. BM amyloid is common in both amyloidosis types, as characterized in an earlier study [70].

Since most biopsies are performed on fat tissue (Sub-Q) in clinical practice in PCD patients, amyloidosis in many other organs, such as the BM, is difficult to detect. Amyloid deposition is present in the BM smears of a proportion of patients affected by multiple myeloma [71]. An earlier study reported that [^11^C]PIB PET/MRI has limitations in the evaluation of fat tissue [72]. In the present study, [^18^F]FBP PET was able to detect fat in 40-55% of patients in the PCD, AL and hATTR-PN groups. [^18^F]FBP is excreted through the hepatobiliary system; therefore, it is physiologically taken up in the urinary tract (kidney, renal pelvis, ureter and bladder) and enterohepatic circulatory system (liver, gallbladder, bile duct and small intestine). The evaluation of [^18^F]FBP in the liver, bladder, etc., might thus have limitations.

[^18^F]FBP SUV and visual analysis provide comparable measures of the number of organs involved in PCD and AL, as well as the hATTR-PN group, and are more sensitive than [18F]FDG PET and clinical evaluation. In comparison, [^18^F]FDG SUV analysis was more sensitive than visual analysis, with a significantly greater degree of organ involvement detected in the AL group. To date, only a case study using [^18^F]FDG and [^18^F]FBP for detecting amyloidosis in the AL has been reported [73]. An earlier meta-analysis across 13 studies (including 90 patients) revealed that the pooled sensitivity of amyloid PET was 0.97, and the pooled specificity was 0.98 [74]. In the present study, [^18^F]FBP showed a similar sensitivity of 1 and specificity of 0.97.

There are several limitations in our study. First, many PCD patients do not undergo biopsy or only undergo sub-Q fat biopsy. Second, PET scans of the brain and body were conducted separately. Total-body PET would be ideal for applying identical settings in this case. Third, [^18^F]FDG PET was not performed for all the AL, and PCD cases, and no in the NC or hATTR-PN patients. In addition, we did not perform a follow-up study on the progression or survival of the patients.

## Conclusions

In conclusion, we demonstrated that [^18^F]FBP PET can be used to qualitatively evaluate organ involvement in patients with PCD, AL and hATTR-PN with high sensitivity and specificity. [^18^F]FBP PET provided more sensitive detection of organ involvement than [^18^F]FDG and clinical assessment. Visual analysis and SUV analysis of [^18^F]FBP PET provide comparable measures for evaluating organ involvement. [^18^F]FBP PET is useful for assisting in the early and accurate detection of organ involvement in amyloidosis, as well as for monitoring treatment effects.

## Statements & Declarations

### Funding

YK received funding from the National Natural Science Foundation of China (No. 82272108), the Natural Science Foundation of Shanghai (No. 22ZR1409200), and the Shanghai Science and Technology Innovation Action Plan Medical Innovation Research Project (23Y11903200). GG received funding from the National Natural Science Foundation of China (No. 82071962). RN acknowledges the Swiss Center for Applied Human Toxicology (SCAHT AP-02).

### Competing Interests

The authors have no relevant financial or non-financial interests to disclose. AR, RN are associate editors for special issues at EJNMMI.

### Author contributions

KY and RN designed the study. KY, BH, FX, JX, YG, and HT conducted patient enrollment and PET imaging evaluation. ZZ, QZ, WW, QW and HZ performed the pathological analysis and biochemical analysis. YG and HT provided ethical applications and infrastructure. KY collected the data and performed PET data analysis. KY, LC, and RN performed the data visualization and statistical analysis. KY and RN wrote the paper. All authors have contributed the editing of the manuscript, read and approved the final manuscript.

### Data availability

The datasets are available from the corresponding authors on reasonable request.

### Ethics approval

This study was approved by the Institutional Ethics Review Board (HIRB) of Huashan Hospital, Fudan University (2022-535; meeting number 2022 M-006; date of approval 26th April 2022). Informed consent was signed by the patients, and the patients agreed that their data would be used for publication.

### Consent to participate

The authors affirm that human research participants provided informed consent for publication of the images.

### Consent to publish

The authors affirm that human research participants provided informed consent for publication of the images.

## Acknowledgments

We acknowledge Prof. Fang Xie, Prof. Chuantao Zuo, Dr. Zhengwei Zhang, PET Center, Huashan Hospital, Fudan University, for the production of [^18^F]florbetapir and [^18^F]FDG and for guidance on FBP-PET image interpretation.

## Notes

### Competing Interest Statement

The authors have declared no competing interest.

