## Supplementary material for "Head-to-head comparison of [^18^F]florbetapir and [^18^F]FDG PET for the early detection of amyloidosis in systemic amyloidosis and plasma cell dyscrasias": Suppl Material

**Supplementary files**

STable 1 Analysis of biochemical markers

|  | PCD | AL | hATTR |
| --- | --- | --- | --- |
| n | 28 | 38 | 8 |
| **Thyroid** |  |  |  |
| Thyroid-stimulating hormone (mIU/L) (0.27-4.2) | 4.4±3.4 | 3.6±3.7 | 1.2±0.2 |
| Triiodothyronine (nmol/L) (1.3-3.1) | 1.3±0.3 | 1.3±0.3 | 1.4±0.2 |
| Thyroxine (nmol/L) 66-181 | 83.4±21.2 | 86.4±21.4 | 93.4±14.9 |
| Free triiodothyronine (pmol/L) 3.1-6.8 | 3.9±0.9 | 3.6±1.0 | 4.3±0.4 |
| Free thyroxine (pmol/L) 12-22 | 14.4±2.1 | 16.1±3.8 | 15.9±2.0 |
| Thyroglobulin antibody (U/ml) <115 | 140.9±452.9 | 22.7±32.8 | 47.2±43.8 |
| Thyroid peroxidase antibody (U/ml) <34 | 41.6±80.9 | 17.7±22.4 | 43.9±33.2 |
| Thyroglobulin (ng/ml) 3.5-7.7 | 14.7±11.6 | 17.1±18.1 | 9.3±7.3 |
| Thyrotropin receptor antibody (IU/L) <1.75 | 0.8±0.0 | 0.8±0.1 | 1.5±1.1 |
| Glycated hemoglobin (HbA1c%) 4-6 | 6.2±1.2 | 5.9±0.8 | 5.4±0.3 |
| Blood sugar (mmol/L) 3.9-5.8 | 6.2±1.8 | 5.9±2.3 | 5.1±0.9 |
| Creatinine (μmol/L) 57-111 | 160.8±209.8 | 110.0±152.1 | 63.3±17.6 |
| Urinary protein (g/24h) <0.15 | 0.6±0.8 | 3.0±5.4 | 0.1±0.0 |
| eGFR(ml/min) MDRD ≥90, | 87.9±43.3 | 103.3±46.9 | 132.1±21.7 |
| eGFR(ml/min) EPI ≥90 | 77.3±34.6 | 89.4±30.0 | 107.7±6.3 |
| Myoglobin (ng/ml ) 25-58 | 75.0±103.0 | 65.0±87.7 | 78.4±64.9 |
| CK-MB mass (ng/ml ) ≤3.61 | 2.2±2.3 | 2.9±2.9 | 6.6±5.5 |
| Systolic blood pressure | 133.1±17.8 | 113.1±16.9 | 110.2±14.3 |
| diastolic blood pressure | 79.4±14.1 | 69.9±9.3 | 70.2±8.1 |
| IgM (g/L) 0.3-2.2 | 1.1±1.4 | 0.6±0.4 | 1.0±0.1 |
| IgG (g/L) 8.6-17.4 | 13.6±8.2 | 11.2±13.8 | 15.4±0.6 |
| IgA (g/L) 1-4.2 | 5.6±12.5 | 2.2±3.0 | 3.7±0.4 |
| κ (g/L) 1.7-3.7 | 2.2±1.5 | 2.1±1.8 | 3.1±0.5 |
| λ (g/L) 0.9-2.1 | 4.0±3.9 | 2.2±3.8 | 1.7±0.2 |
| κ/λ 1.35-2.65 | 1.9±3.6 | 2.4±5.4 | 1.8±0.1 |
| β2 microglobulin (mg/l) 0.7-1.8 | 5.0±5.4 | 4.1±3.0 | 1.9±0.4 |
| Urinary -κ- light chain (mg/L) <7.5 | 120.0±307.4 | 56.4±89.5 | 10.7±5.1 |
| Urinary-λ-light chain (mg/L) <4.1 | 85.6±300.6 | 151.5±641.9 | 3.9±0.0 |
| Urinary β2 microglobulin (mg/L) <0.25 | 7.3±22.6 | 3.4±6.0 | 2.7±1.3 |
| Urinary κ/λ 0.7-4.5 | 26.0±56.0 | 7.2±16.4 | 0.2±0.0 |
| IgG- λ % negtaive | 15.4 | 37.5 | 100 |
| Bence-Jones λ % negtaive | 66.7 | 58.1 | 100 |
| Serum amyloid A，SAA（mg/L）＜10 | 35.8 | 39.3 | 4.8 |

**STable 2 Association between [^18^F]FBP and [^18^F]FDG visual (v) and SUV vs. Clinical diagnosis (Dx) assessment**

|  | NC | PA* | AL | hATTR | **PA vs. NC** | **AL vs. NC** | **hATTR vs. NC** | **AL vs. PA** |
| --- | --- | --- | --- | --- | --- | --- | --- | --- |
| [^18^F]FBP SUV (n) | 0.4±1.1(8) | 2.5±1.4(26) | 4.1±2.3(38) | 3.9±1.0(8) | 0.0062 | <0.0001 | <0.0001 | <0.0001 |
| [^18^F]FDG SUV(n) |  | 2.1±1.5(16) | 2.8±1.8(26) |  |  |  |  | ns |
| [^18^F]FBP visual (n) | 0.4±0.7(8) | 2.6±1.6(28) | 4.0±2.0(38) | 3.8±1.4(8) | 0.0024 | <0.0001 | 0.0002 | 0.0049 |
| [^18^F]FDG visual (n) |  | 1.5±1.5(15) | 1.1±1.0(26) |  |  |  |  | ns |
| Clinical (Dx) | 0.3±0.7(8) | 0.0±0.2(25) | 2.3±1.3(36) | 1.6±0.7(8) | ns | 0.0059 | ns | <0.0001 |
| [^18^F]FBP SUV vs.Dx | ns | <0.0001 | <0.0001 | 0.0139 |  |  |  |  |
| [^18^F]FBP visual vs. Dx | ns | <0.0001 | 0.0002 | 0.0223 |  |  |  |  |
| [^18^F]FDG SUV vs. Dx |  | <0.0001 | ns |  |  |  |  |  |
| [^18^F]FDG v vs. Dx |  | <0.0001 | ns |  |  |  |  |  |
| [^18^F]FBP SUV vs. [^18^F]FDG SUV |  | 0.0008 | 0.0301 |  |  |  |  |  |
| [^18^F]FBP SUV vs. [^18^F]FBP v |  | ns | ns |  |  |  |  |  |
| [^18^F]FBP SUV vs. [^18^F]FDG v |  | ns | <0.0001 |  |  |  |  |  |
| [^18^F]FDG SUV vs. [^18^F]FBP v | ns | ns | ns | ns |  |  |  |  |
| [^18^F]FDG SUV vs. [^18^F]FDG v |  | ns | 0.0102 |  |  |  |  |  |
| [^18^F]FBP v vs. [^18^F]FDG v |  | ns | <0.0001 |  |  |  |  |  |

STable 3 Correlation of [^18^F]**FBP** and [^18^F]FDG in each organs in AL and PCD cases

| **SUV mean** | [^18^F]**FBP** | [^18^F]FDG | p | r |
| --- | --- | --- | --- | --- |
| parotid gland | 2.492 | 1.584 | ns |  |
| tongue | 1.785 | 1.454 | ns |  |
| submandibular gland | 1.567 | 2.065 | ns |  |
| thyroid | 1.432 | 1.527 | ns |  |
| humeral head | 1.184 | 0.8892 | 6.48E-07 | 0.7376 |
| humerus | 1.967 | 1.341 | 0.0006 | 0.6465 |
| muscle | 1.333 | 0.9324 | 0.0021 | 0.5079 |
| lymph nodes | 1.247 | 1.73 | ns |  |
| lung | 0.856 | 0.6568 | 2.06E-07 | 0.7581 |
| heart | 2.413 | 2.205 | ns |  |
| mediastinal blood pool | 0.5987 | 1.324 | 0.0002 | 0.5979 |
| liver | 8.207 | 2.327 | ns |  |
| stomach | 1.737 | 1.251 | 0.0015 | 0.5220 |
| spleen | 1.276 | 2.086 | 0.0398 | 0.3543 |
| pancreas | 1.715 | 1.77 | 0.0183 | 0.4021 |
| kidney | 1.945 | 2.462 | ns |  |
| intestinal | 0.9333 | 0.8541 | 0.0182 | 0.4025 |
| fat | 1.099 | 0.5459 | 0.0002 | 0.5930 |
| bone marrow | 3.229 | 2.4 | ns |  |

Nonparametric Spearman’s rank analysis

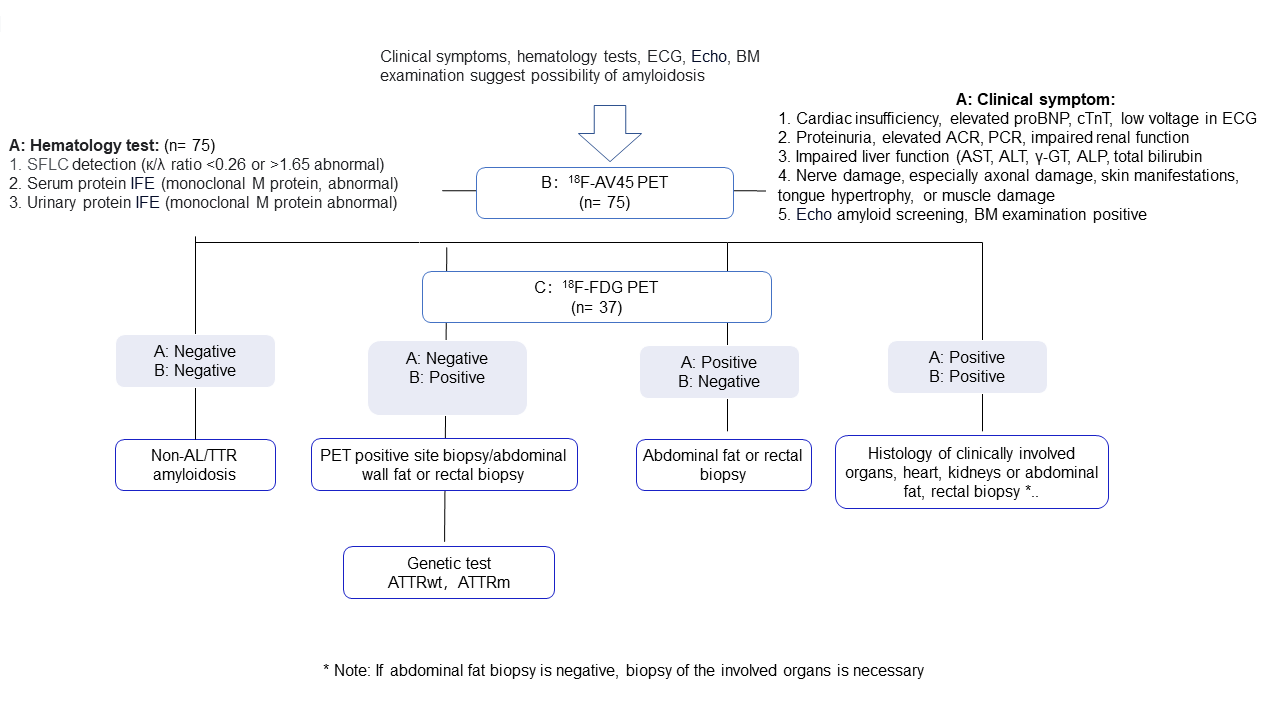
**SFig. 1 Clinical workflow**. * Note: If histology of abdominal fat biopsy is negative, biopsy of the involved organs is necessary. BM, bone marrow; Echo, Cardiac Ultrasound; ECG, Electrocardiogram; SFLC, Serum free light chain; IFE, immunofixation electrophoresis

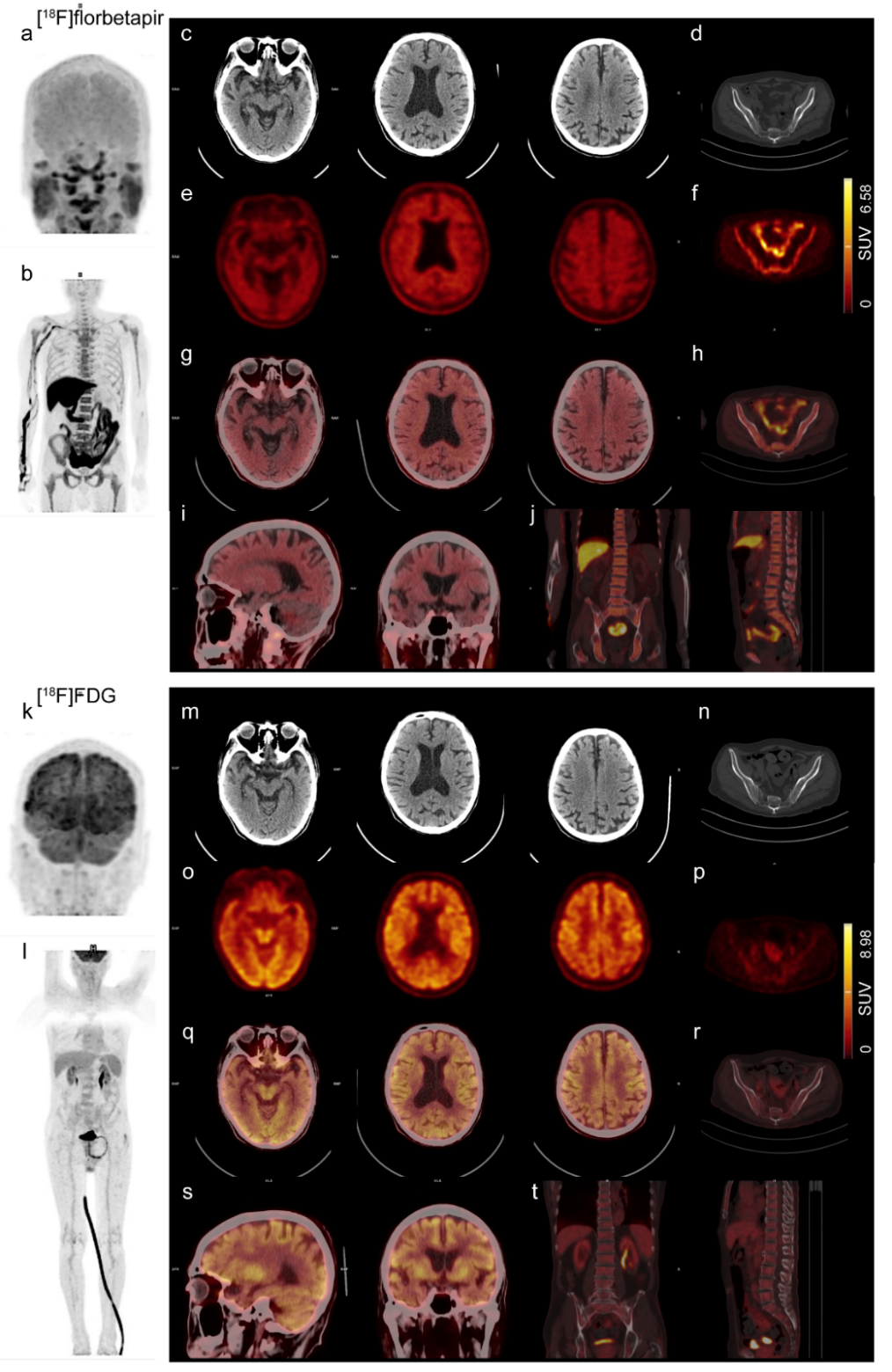

**SFig 2. Increased uptake of [^18^F]florbetapir (FBP) and [^18^F]FDG PET in the brain and spinal cord of one patient with MM.** Age 83 years old, male. The patient is negative for AL and negative in the [^18^F]FBP (a-j) and [^18^F]FDG (k-t) uptake in the peripheral. 3D MIP in the brain (a, k) and in the spinal cord (b, l). Transaxial view of CT (a, m), PET (e, o), transaxial, sagittal and coronal views of PET/CT overlay images of the brain (g, i, q, s) view; Transaxial view of CT (d, n), PET (f, p), transaxial, sagittal and coronal (h, j, r, t) view of PET/CT overlay images of the spinal cord.

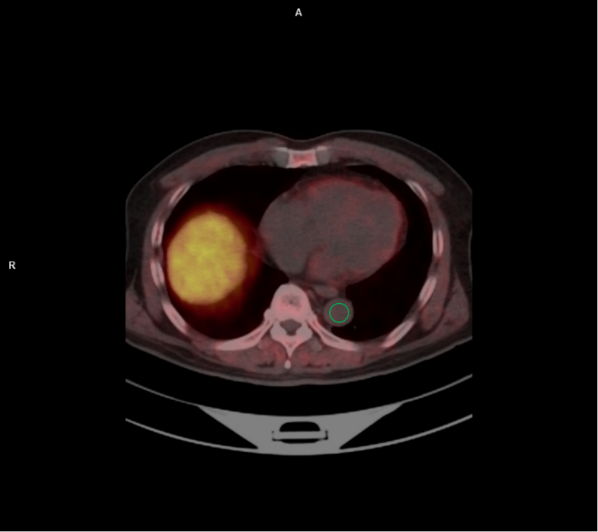

**SFig. 3 Reference region for TBR analysis of [^18^F]FBP and [^18^F]FDG PET**. Mediastinal blood pool ROI diameter, diameter size 1 cm (Green circle).

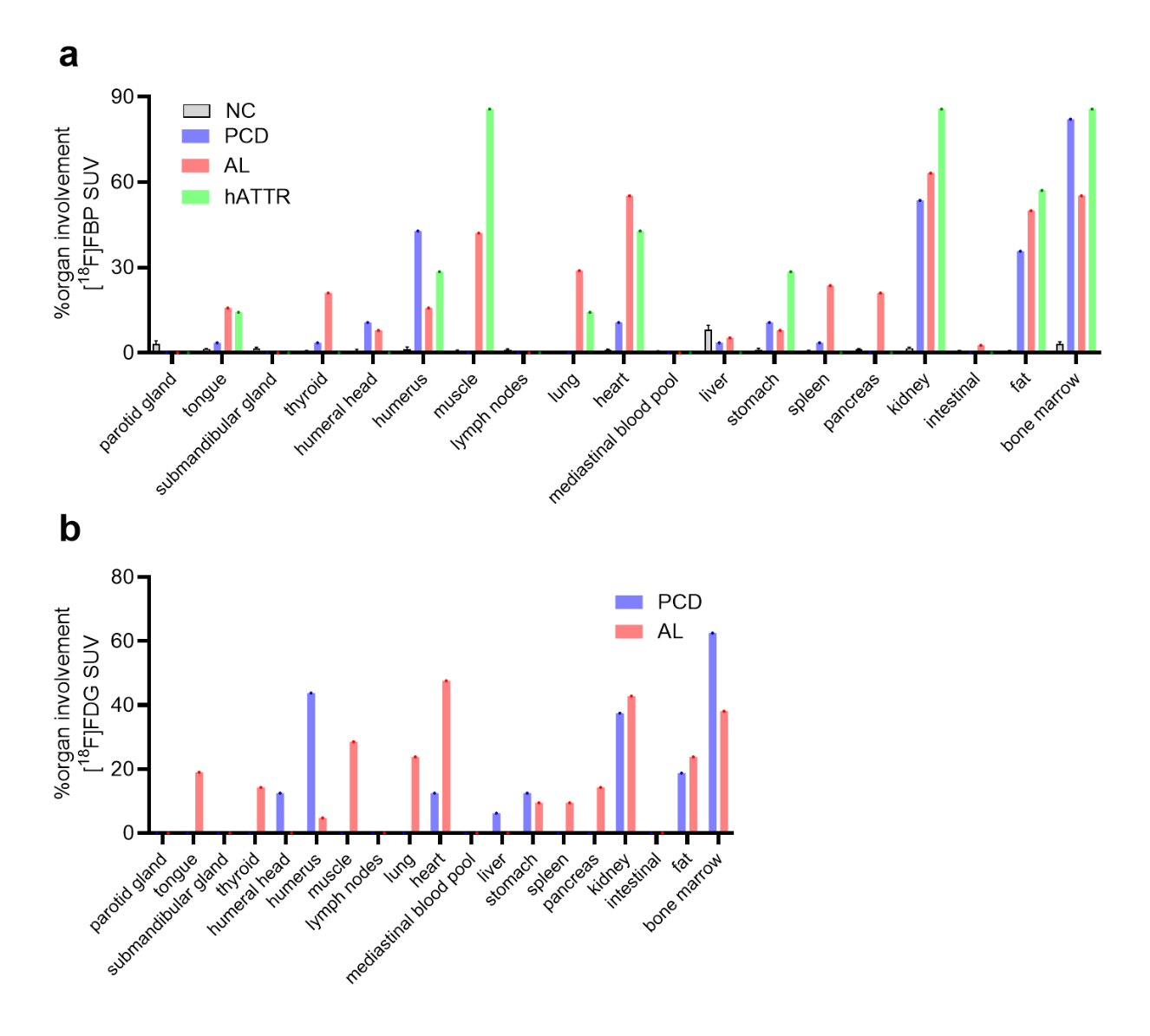

**SFig 4** **Evaluation of % organ involvement by[^18^F]FBP and [^18^F]FDG PET.** (a) [^18^F]FBP SUV analysis in NC, PCD, AL and hATTR patients; (b) [^18^F]FDG PET SUV analysis in PCD, AL patients;

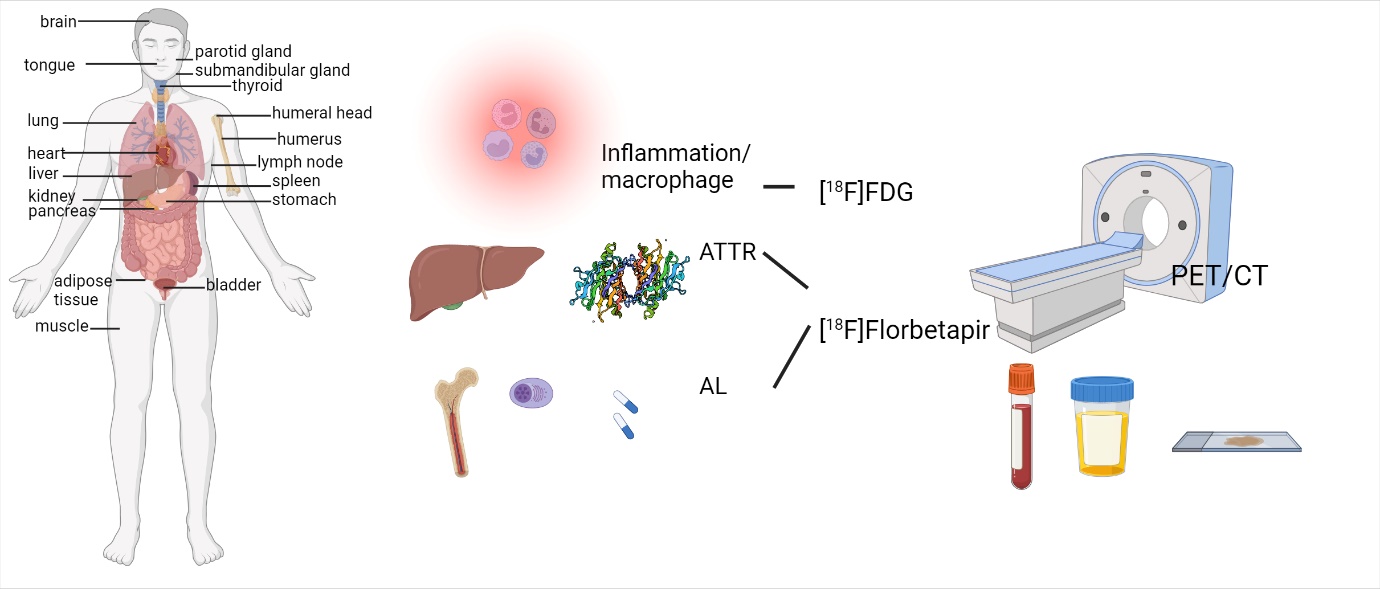

Graphic abstract
